# Neonatal Presentations to the Children’s Emergency Department

**DOI:** 10.1101/2020.09.07.20190140

**Authors:** Sarah Blakey, Mark D Lyttle, Dan Magnus

**Author notes:** Corresponding author:* Dan Magnus: Children’s Emergency Department, Bristol Royal Hospital for Children, Upper Maudlin Street, Bristol, BS2 8BJ.

## Abstract

**Background:** Paediatric attendances to Emergency Departments (EDs) in the UK are increasing, particularly for younger children. Neonates present a challenge due to their non-specific presentations. Community services are under increasing pressure and parents may preferentially bring their children to the ED, even for non-urgent problems. Neonatal attendances have not been extensively studied, but previous reviews have shown many are well, often not requiring specific medical intervention. This study aimed to characterise the presenting features, management and disposition of neonatal attendances to a tertiary Children’s ED (CED).

**Methods:** Retrospective observational review of medical records identified via the ED Electronic database of neonatal attendances (≤28 days) to Bristol Royal Hospital for Children (BRHC) over 12 months (01/01/2016-31/12/2016). Further information was obtained from investigation results, discharge summaries and historical admissions data.

**Results:** Neonatal attendances increased from 655 to 1205 from 2008-2016. The most common presenting complaints were breathing difficulty (18.1%), vomiting (8.3%) and poor feeding (8.2%). The most common diagnoses were ‘no significant medical problem’ (41.9%), bronchiolitis (10.5%) and suspected sepsis (10.0%). Just over 1/3 were admitted (23% inpatient, 12% Short Stay Unit). Median length of stay for inpatients was 2 days. Half of neonatal attendances to the ED had no investigations performed and most (77.7%) needed advice or observation only.

**Conclusion:** Many neonates presenting to the CED were well and discharged with observation only. This suggests not only that there is potential for improved community management but that increased support for community colleagues and new parents is needed. There are also implications for reviewing training in emergency medicine, especially the ability to assess ‘well’ infants and to manage common neonatal problems. Drivers of health policy should consider developing enhanced models of out of hospital care which are acceptable to clinicians and families

## INTRODUCTION

Emergency Departments (EDs) are under increasing pressure, with rising attendances and admission rates. In the UK there has been a 14% increase in childhood emergency admissions over the past decade, a rate which is doubled in infants under one year old. (1) One potentially modifiable pattern is short term admissions for minor conditions which may be managed away from hospital. (2) However community services are also under pressure. Even though women would like more contact with postnatal community practitioners, Health Visitor (HV) numbers have dropped by 20% (3). This, combined with earlier postnatal discharge, contributes to increased community case load (4), with consequent rises in ED attendances (5) for conditions traditionally managed by other health care professionals. (6)

ED attendances by neonates (≤28 days old) are rising disproportionately quicker than older infants. (5) However most studies to date are limited by small numbers, and their applicability to our setting is unknown. (7) In Europe (8-10) and North America (5,11), up to one-third of neonates attending EDs have no medical issue, and a high proportion have low acuity problems requiring no medical investigation or treatment. Parents prefer to bring their children to either general(12) or paediatric (13) EDs, even with non-urgent problems, for reasons including parental anxiety, perceived advantages of EDs (resources and expertise), and convenience.

Clinical assessment of neonates is challenging. They are a vulnerable and high-risk cohort, who exhibit non-specific symptoms even with serious illnesses.(14) Risk tolerance amongst non-hospital healthcare providers is therefore understandably low, given the risk of rapid deterioration and an inability to send and act on investigations. Although EDs have access to resources to exclude serious conditions, they are suboptimal environments for assessing well neonates due to increased exposure to infections, and variable training for ED staff in normal newborn problems, leading to potential over-investigation and treatment.

Reducing ED attendances for neonates with benign conditions may benefit patients, families, and healthcare systems. The low proportion of neonates requiring medical interventions seen in other settings suggests alternative clinical pathways may be appropriate in our setting if these findings are replicated. We therefore aimed to evaluate the characteristics, referral source and disposition of neonatal ED attendances in our institution.

## METHODS

This retrospective chart review study evaluated attendances to a tertiary urban children’s ED of all neonates (≤28 days of age) between 1^st^ January and 31^st^ December 2016, and is reported in accordance with the RECORD statement. There were no exclusion criteria. This ED provides local secondary level emergency care and is the regional tertiary centre for medical and surgical specialties, and paediatric major trauma. The hospital operates a “single front door” system, with the ED receiving all emergency attendances including referrals to inpatient specialty teams.

Participants were identified using electronic patient tracking systems. Data abstracted included sex, age, time/day of attendance, triage category, disposition, presenting complaint, diagnosis, investigations, and treatment. Admission was defined as a stay of any duration in any inpatient area including the short stay unit (SSU). Presenting complaints and diagnoses were categorised by one investigator (SB), with clarification sought from a second investigator (DM) where the category was unclear. Presenting complaints were verbatim information given by parents on ED arrival. We defined the diagnostic category “no significant medical problem” as “well babies with no identified pathology after assessment, with or without investigations or a period of observation/admission”. “Suspected sepsis” was ascribed as a diagnosis when a clinician described this in their impression and instigated invasive tests to rule sepsis in/out, and started intravenous antibiotic treatment.

We used a convenience sample of attendances over a one-year period to account for any seasonal variation. Data were classed as missing if they were unavailable on medical chart review. Data were analysed using STATA V. 12.0 (15) and results are presented using descriptive statistics, utilising proportions for discrete variables, and means with standard deviations where appropriate. No tests for association of statistical significance were deemed appropriate given the study design. Attendance and admission data from 2008-2016 were reviewed to determine whether overall attendance rates in our institution were increasing in line with previous studies.

This study was approved as a service evaluation using routinely collected, existing, anonymised data by University Hospitals Bristol NHS Foundation Trust following assessment against the Health Research Authority Framework. (16) Raw data is available on request from study authors.

## RESULTS

Overall ED attendances rose by 41% from 29,164 in 2008 to 41,067 in 2016, with a disproportionate rise in neonatal attendances from 655 to 1205 (84% increase). Neonatal admissions increased from 262 to 489 over this period, though the proportion of patients admitted remained constant (40% in 2008; 41% in 2016).

The 1205 attendances in 2016, by 1104 neonates, accounted for 3% of all ED attendances, and represented approximately 8% of babies born in the catchment area (Figure 1). Data were missing for referral source in two cases, presenting complaint in one case and disposition in 6 cases; data were otherwise complete. Mean age was 14 days (SD +/− 8 days), and 679 (56%) were male; nearly half were brought directly by parents and one-third were referred by community colleagues. (Table 1) Nearly two-thirds of attendances were discharged home from the ED (754, 63%). (Table 2)

**Figure 1:**
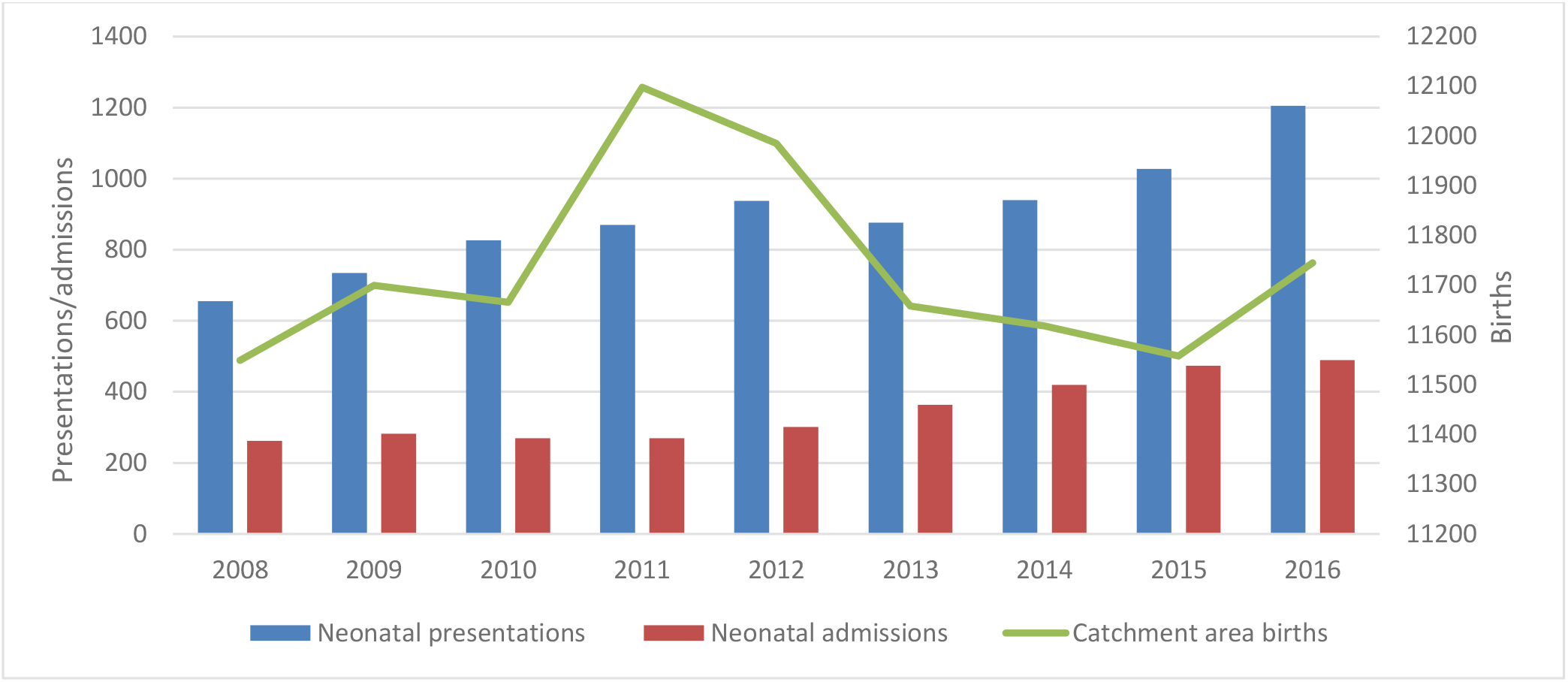
Neonatal attendance and admission rates from 2008–2016 with Catchment Area Births.

**Table 1:** Demographics, and ten most common presenting complaints and diagnoses.

|  | Frequency (%) of attendances |
| --- | --- |
| Male | 679 (56.4) |
| Female | 526 (43.6) |
| *Age (Mean 14 days; SD +/- 8 days)* |  |
| 0-7 days | 326 (27.0) |
| 8-14 days | 347 (28.8) |
| 15-21 days | 280 (23.2) |
| 22-28 days | 252 (20.9) |
| Secondary care level ED attendance | 1049 (87.1) |
| Referred for tertiary opinion/investigation | 156 (12.9) |
| *Day of Attendance* |  |
| Weekday | 882 (73.2) |
| Weekend | 323 (26.8) |
| *Time of attendance* |  |
| 07:00-11:59 | 146 (12.1) |
| 12:00-17:59 | 440 (36.5) |
| 18:00-22:59 | 347 (28.8) |
| 23:00-06:59 | 272 (22.6) |
| *Referral Method* |  |
| Self | 584 (48.5) |
| GP | 255 (21.2) |
| Midwife | 155 (12.9) |
| Health Visitor | 18 (1.5) |
| Transfer | 158 (13.1) |
| Asked to attend by speciality team | 33 (2.7) |
| *Presenting Complaint* |  |
| Breathing Difficulty | 218 (18.1) |
| Vomiting | 100 (8.3) |
| Poor Feeding | 99 (8.2) |
| Contrast Study | 89 (7.4) |
| Fever | 71 (5.9) |
| Specialty Admission | 62 (5.2) |
| Unsettled | 61 (5.1) |
| Unwell | 53 (4.4) |
| Rash/Skin Problem | 52 (4.3) |
| Jaundice | 44 (3.7) |
| *Diagnosis* |  |
| No significant medical problem | 504 (41.9) |
| Bronchiolitis | 126 (10.5) |
| Sepsis (Suspected or confirmed) | 121 (10.0) |
| GORD | 74 (6.2) |
| Surgical Problem | 67 (5.6) |
| Upper Respiratory Tract Infection | 48 (4.0) |
| Local Infection | 26 (2.2) |
| Head Injury (minor) | 24 (2.0) |
| Constipation | 23 (1.8) |
| Birth Injury | 20 (1.7) |

SD: Standard deviation; GORD: Gastro-oesophageal Reflux Disease

**Table 2:** Diagnoses, disposition of patients, and proportions requiring no investigations by community referral source.

|  | Whole Cohort<br>(N = 1205)<br>n (%) | GP<br>(N=255)<br>n (%) | Health Visitor<br>(N=18)<br>n (%) | Midwife<br>(N=155)<br>n (%) | Self<br>(N = 584)<br>n (%) |
| --- | --- | --- | --- | --- | --- |
| *Common Diagnoses* |  |  |  |  |  |
| No significant medical problem | 504 (41.8) | 82 (32.2) | 10 (55.6) | 84 (54.1) | 255 (43.7) |
| Bronchiolitis | 126 (10.5) | 35 (13.7) | 1 (5.6) | 4 (2.6) | 84 (14.4) |
| Suspected Sepsis | 121 (10.0) | 27 (10.6) | 0 (0.0) | 16 (10.3) | 62 (10.6) |
| GORD | 74 (6.1) | 29 (11.4) | 4 (22.2) | 6 (3.4) | 22 (3.8) |
| URTI | 51 (4.2) | 15 (5.9) | 1 (5.6) | 3 (1.9) | 32 (5.5) |
| *Disposition* |  |  |  |  |  |
| Discharged | 754 (63.0) | 162 (63.5) | 15 (83.3) | 106 (68.3) | 366 (62.6) |
| SSU | 155 (12.9) | 28 (10.9) | 2 (11.1) | 15 (9.7) | 101 (17.3) |
| Ward | 280 (23.4) | 63 (24.7) | 1 (5.6) | 34 (21.9) | 110 (18.8) |
| PICU | 10 (0.8) | 2 (0.8) | 0 (0.0) | 0 (0.0) | 7 (1.2) |
| Proportion requiring no investigation | 601 (49.9) | 132 (51.7) | 14 (77.8) | 59 (38.1) | 330 (56.5) |

*GORD: Gastro-oesophageal Reflux Disease; URTI: Upper Respiratory Tract Infection; SSU: Short Stay Unit; PICU: Paediatric Intensive Care Unit*

“Breathing difficulty” was the most common presenting complaint (218/1205; 18%); vomiting and poor feeding were the next most common (8% each). The most common diagnosis was “no significant medical problem” (504/1205; 42%). Common acute pathologies included bronchiolitis (126/1205; 11%), and suspected sepsis (121/1205; 10%). Common diagnoses for each referral source are shown in Table 2.

For neonates discharged directly from the ED, over half (411, 54.5%) had “no significant medical problem”. (Table 3) For those admitted to inpatient wards, suspected sepsis was the most common diagnosis. Of the 106 (88%) septic screens available for review, five neonates had invasive bacterial infection (IBI; four positive blood cultures, three positive cerebrospinal fluid (CSF) culture). Nineteen patients had a positive urine culture (17 E. Coli), and Enterovirus was detected in the CSF of 21 (20%) patients.

**Table 3:** Common diagnoses, and proportions requiring investigation and treatments categorised by disposition.

|  | Discharged (n=754)<br>N (%) |  | Short Stay Unit<br>(n=155)<br>N (%) |  | Inpatient Ward<br>(n=280)<br>N (%) |  | PICU (n=10)<br>N (%) |  |
| --- | --- | --- | --- | --- | --- | --- | --- | --- |
| **Top 5 Diagnoses** | No significant medical problem | 411 (54) | No significant medical problem | 76 (51) | Suspected sepsis | 105 (38) | Suspected sepsis | 5 (50) |
|  | Bronchiolitis | 58 (8) | Bronchiolitis | 27 (18) | Surgical problem | 51 (18) | Bronchiolitis | 2 (20) |
|  | GORD | 53 (7) | URTI | 14 (9) | Bronchiolitis | 35 (13) | Metabolic | 1 (10) |
|  | URTI | 31 (4) | Reflux | 13 (9) | No significant medical problem | 17 (6) | Seizures | 1 (10) |
|  | Constipation | 20 (3) | Gastroenteritis | 5 (3) | Congenital Heart Disease | 9 (3) | SVT | 1 (10) |
| **Investigations performed\*** | 283 (37) |  | 104 (67) |  | 207 (74) |  | 10 (100) |  |
| **Treatment given\*** | 79 (10) |  | 15 (10) |  | 165 (59) |  | 10 (100) |  |

*\*Refers to number of patients, not number of individual investigations or treatments; PICU: Paediatric Intensive care unit; URTI: Upper Respiratory Tract Infection; SVT: Supraventricular Tachycardia; GORD: Gastro-oesophageal Reflux Disease*

Half (601/1205) had no laboratory investigations performed. The most common investigations were blood tests (305; 25%), blood gases (261; 22%), urine samples (218; 18%) and lumbar punctures (LP) (97; 8%). Investigations were less frequently performed in those discharged directly from the ED (37%) than those admitted to the SSU (68%) and inpatient wards (74%) (Table 3). Proportions requiring investigations categorised by referral source are shown in Table 2.

Most families (936, 78%) required advice or reassurance only, and no pharmacological treatment. Patients discharged directly or admitted to SSU had treatment commenced in 10% of cases; of those admitted to inpatient wards, treatment was commenced in 59% (Table 3)

Greater proportions of attendees with no significant medical problem were < 14 days, and had a triage category of 3 or 4, compared to those with another diagnosis. (Table 4) Proportions with no investigations were similar, but those with another diagnosis were more likely to have had multiple investigations (14.9 vs 27.3%).

**Table 4:**
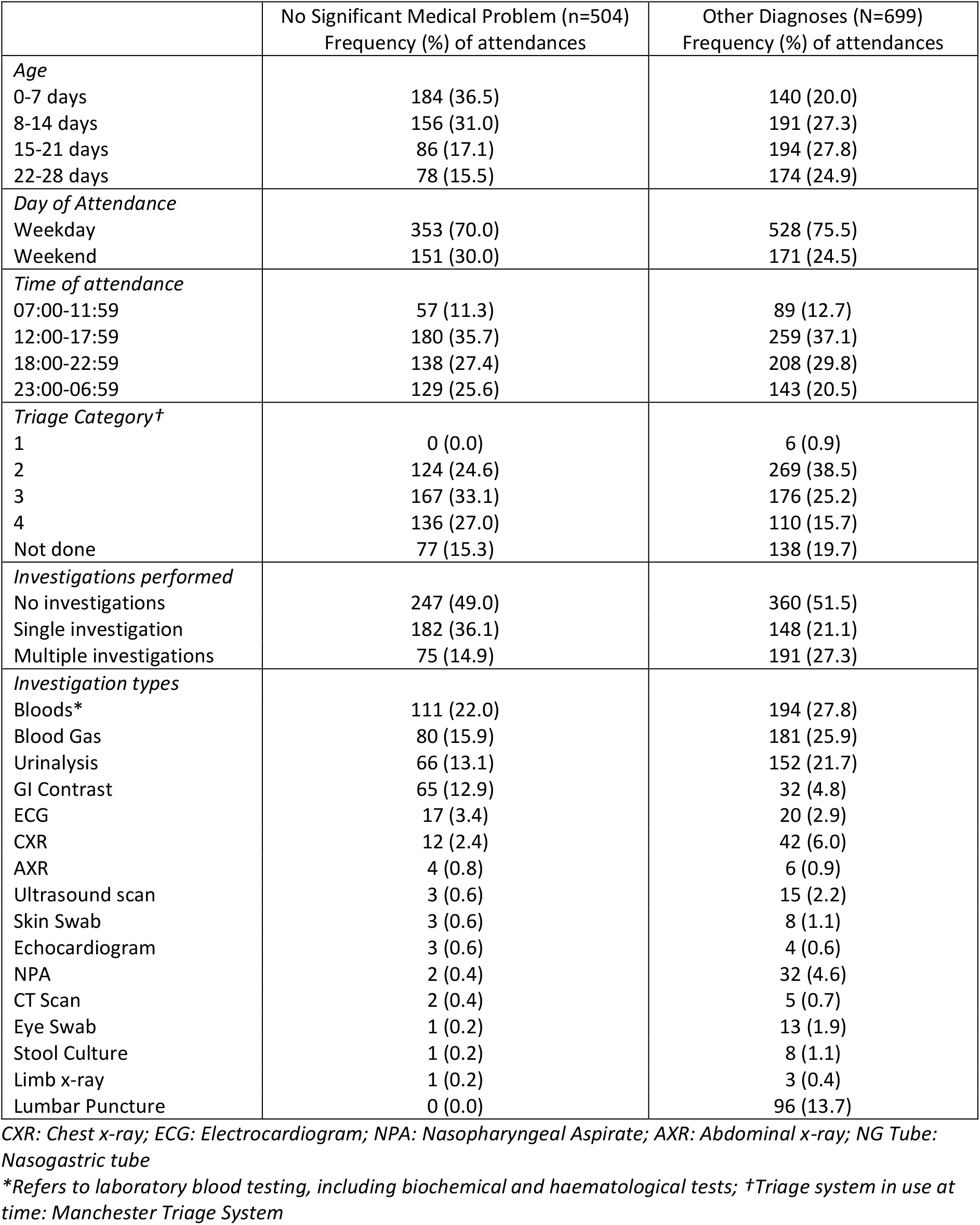
Comparison of patients diagnosed as “no significant medical problem” against those with other diagnoses.

Ninety five (9%) patients had at least one re-attendance before the age of 28 days, of which 34 (36%) had unrelated presenting complaints; the most common diagnosis when re-attending for the same problem was bronchiolitis (n = 12). Thirteen were planned re-attendances for repeat investigations or medical review. Sixteen of those with an initial diagnosis of “no significant medical problem” returned with a similar presenting problem and were discharged with no change in diagnosis; five had a change in diagnosis (two with bronchiolitis, one with constipation, one with gastroenteritis, and one with gastro-oesophageal reflux disease).

## DISCUSSION

In this chart review study, many neonates attending our ED were well babies, with the most common diagnosis being “no significant medical problem”. Half were brought directly to the ED by parents, two-thirds were discharged directly home, half had no investigations, and most did not need pharmacologic treatment. Common pathologies requiring admission included bronchiolitis and suspected sepsis, but rates of proven IBI were low.

In keeping with other sources, (5) the rise in neonatal presentations over an eight year period was disproportionate to the rise in all-cause attendances, though the proportion requiring admission remained constant. Half were self-referrals (of which only one-fifth required inpatient admission), and over half had no investigations. This combination, along with a static birth rate, gives rise to the possibility that parental fears and expectations may be key drivers, as reflected in one recent study wherein parents felt a same day clinical review was warranted even if their baby’s condition was not thought to be serious (17). A short period of observation provides reassurance, but for several reasons may not be sub-optimal, including pathogen exposure and medicalisation of normal neonatal behaviours. Support and education for new parents highlighting these potential issues, while taking into account factors such as media-induced fear and access to primary care, are likely to be crucial interventions if this trend is to be reversed (18). Capitalising on technological advances may be of benefit, with one internet-based home monitoring system resulting in reduced neonatal ED attendances, (19) but such community based interventions may not always have the expected impact if attitudes towards ED use are not considered (20).

Any policy aimed at appropriate use of non-hospital healthcare resource should focus on some key patient cohorts, and ensure the model of care is feasible and acceptable. A larger proportion of neonates in our study were “well babies” when compared to European cohorts (9,21) though this may reflect literature-based or clinician differences in defining pathology. In our study, those with no significant medical problem were more likely to be < 14 days old, had a lower triage category, and were less likely to have multiple investigations than those with pathology. An additional cohort were diagnosed with non-emergency problems such as reflux and constipation, and common acute diagnoses included bronchiolitis and URTIs, conditions which may be manageable in community settings. However over two thirds of patients referred from other healthcare professionals were discharged home from the ED, many without investigations. Models of care which provide appropriate community-based resource and expertise can not only reduce hospital attendances, but also strengthen primary/secondary care relationships and increase knowledge. The Child Health Hub Model (22) is an example of a model which may bring benefits in the short and long term, by making expertise available at the point of care in the community, and by disseminating expertise to other healthcare professionals. Equally, collaborative work with families, identifying essential characteristics that would make community services the preferred option for neonatal issues, is essential to inform any such interventions.

Admission rates in our study were higher than in North America (5) but similar or lower than Europe. (7,8) Such differences may be multifactorial and reflect system/reporting differences, practice variation, or patient characteristics, but it is not possible to robustly interrogate these factors given the level of detail available in other reports. For example, direct admissions to specialty teams are included in our ED workload, the 4 hour target for completion of care in the UK influences admission rates, and definitions of admission vary (in our study, SSU admissions were included which accounted for one-third of all admissions).

For neonates admitted to inpatient wards, suspected sepsis was the most common presumptive diagnosis, though the prevalence of IBI (positive blood or CSF culture) was only 4.7% in those screened for sepsis. Previous reviews of neonatal ED attendances have not given details of culture results, but rates of infection were similar to other literature (14) though this also varies depending on the cohort studied (< 28days vs. < 90days) and whether UTIs are included (IBI vs, non-IBI).

NICE guidelines (23) have a low threshold for investigation of febrile neonates, suggesting those presenting with a fever (≥38®C) have a full septic screen, including CSF culture. Research is ongoing, focusing on the development of prediction tools to identify infants at low risk of IBI (24), both with and without results of investigations as predictor variables (25). Tools which include inflammatory markers (particularly Point of Care Tests; POCT) may reduce neonatal attendances and admissions, as these could be rapidly completed and interpreted both in primary and secondary care. This same principle could be applied to a number of conditions for which management pathways follow test result thresholds (eg jaundice, weight loss), but there is to date little published research such applications of POCT in neonates.

To our knowledge, this is the largest European review of neonatal presentations to a CED. The main limitation is the single-centre retrospective observational design. Retrospective chart review relies on good coding and documentation which may not always be present. However we selected variables more likely to be completed more fully and accurately, as they are contemporaneously captured on electronic systems. A single clinician reviewed the majority of medical notes and this type of review is recognised to have potential for misclassification bias. All categorisations and definitions were therefore agreed a priori by consensus of the study team, with a route to second reviewer clearly identified. As a tertiary centre, the findings from this study may not be generalisable to other settings, but the function of the ED in providing secondary level care to a wide mixed urban and semi-rural catchment area mitigates this. In addition, provision of community neonatal services may be different in our regions compared to other regions, though currently policy reports and literature reflect similar challenges and changes on a national level.

In conclusion, whilst neonatal attendances to our CED are increasing disproportionately to other cohorts, large proportions are clinically well and discharged home without investigation. Policy makers should consider implementing collaborative models of infant healthcare outside hospitals, which may reduce ED neonatal attendances if feasible to deliver, and acceptable to families. Further focus should be on the development of safe, evidence based prediction tools which support clinician decision making in primary and secondary level care.

## Data Availability

Raw data available from authors on request

